# The Medicines Intelligence Data Platform: A population-based data resource from New South Wales, Australia

**DOI:** 10.1101/2024.04.29.24306520

**Authors:** Helga Zoega, Michael O Falster, Malcolm B Gillies, Melisa Litchfield, Ximena Camacho, Claudia Bruno, Benjamin Daniels, Natasha Donnolley, Alys Havard, Andrea L Schaffer, Georgina Chambers, Louisa Degenhardt, Timothy Dobbins, Natasa Gisev, Rebecca Ivers, Louisa Jorm, Bette Liu, Claire M Vajdic, Sallie-Anne Pearson

**Author notes:** **Correspondence:** Professor Sallie-Anne Pearson, Director, NHMRC Medicines Intelligence Centre of Research Excellence, School of Population Health, Faculty of Medicine and Health, Rm 230, Samuels Building, UNSW SYDNEY NSW 2052 AUSTRALIA, E. joint first authors (Helga Zoega, Michael O Falster).

## Abstract

The *Medicines Intelligence (MedIntel) Data Platform* is an anonymised linked data resource designed to generate real-world evidence on prescribed medicine use, safety, costs and cost-effectiveness in Australia. The platform comprises Medicare-eligible people who are ≥18 years and residing in New South Wales (NSW), Australia, any time during 2005-2020, with linked data on dispensed prescription medicines (Pharmaceutical Benefits Scheme), health service use (Medicare Benefits Schedule), emergency department visits (NSW Emergency Department Data Collection), hospitalisations (NSW Admitted Patient Data Collection), cancer notifications (NSW Cancer Registry), fact and cause of death (National Death Index). Data are currently available to 2022, with approval to update the cohort and data collections annually.

The platform includes 7.4 million unique people across all years, covering 36.9% of the Australian adult population; the overall population increased from 4.8M in 2005 to 6.0M in 2020. As of 1 January 2019 (the last pre-pandemic year), the cohort had a mean age of 48.7 years (51.1% female), with most people (4.4M, 74.7%) residing in a major city. In 2019, 4.4M people (73.3%) were dispensed a medicine, 1.2M (20.5%) were hospitalised, 5.3M (89.4%) had a GP or specialist appointment, and 54 003 people died. Anti-infectives were the most prevalent medicines dispensed to the cohort in 2019 (43.1%), followed by nervous system (32.2%) and cardiovascular system medicines (30.2%).

The *MedIntel Data Platform* creates opportunities for national and international research collaborations and enables us to address contemporary clinically- and policy-relevant research questions about quality use of medicines and health outcomes in Australia and globally.

**Five Key Points:**

1. The *Medicines Intelligence (MedIntel) Data Platform* is a purpose-built, real-world data resource designed to generate evidence on prescribed medicine use, safety, costs and cost-effectiveness in Australia
2. This is a whole of population anonymised linkage of medicine and health records of Medicare-eligible adults (≥18 years) residing in Australia’s most populous state, New South Wales, from 2005 onwards
3. The platform comprised 7.4M unique people across all years, representing 36.9% of the Australian adult population; the population increased from 4.8M in 2005 to 6.0M in 2020
4. As of 1 January 2019 (the last pre-pandemic year), the cohort had a mean age of 48.7 years (51.1% female), with most people (4.4M, 74.7%) residing in a major city
5. The *MedIntel Data Platform* creates unique research opportunities for national and international research collaborations exploring population-level medicine use and outcomes

**Plain Language Summary:** The *Medicines Intelligence (MedIntel) Data Platform* is a new linked data resource established to generate evidence on prescribed medicine use, safety, costs, and cost-effectiveness in Australia. It adheres to best practice privacy principles, with no identifying information available to researchers. The platform comprises Medicare-eligible people who are ≥18 years and residing in New South Wales (NSW), Australia, any time during 2005-2020, with linked data on dispensed prescription medicines, Medicare services, emergency department visits, hospitalisations, cancer notifications, and deaths. In total, the platform includes 7.4 million unique people across all years, covering 36.9% of the Australian adult population. As of 1 January 2019 (the last pre-pandemic year), the cohort had a mean age of 48.7 years (51.1% female), with most people (4.4M, 74.7%) residing in a major city. In 2019, 4.4M people (73.3%) were dispensed a medicine (most commonly anti-infective, nervous system, and cardiovascular medicines), 1.2M (20.5%) were hospitalised, 5.3M (89.4%) had a GP or specialist appointment, and 54 003 people died. Data are available until 2022 with approval for annual updates. This platform creates opportunities for national and international research collaborations and enables us to address important questions about quality use of medicines and health outcomes.

## Introduction

Insights generated from routinely collected health and social data have enormous potential to improve the use, effectiveness, safety, costs and cost-effectiveness of prescribed medicines. Data resources of this kind, make it possible to study the outcomes of medicine use in everyday clinical practice and among populations rarely included in randomised clinical trials.^1^

Due to its universal healthcare system, Australia has accumulated some of the world’s most comprehensive health data assets,^2,3^ including medicines dispensed under its national subsidy program, the Pharmaceutical Benefits Scheme (PBS), and hospital records collected by the State and Territory governments. Despite this, Australia has only partially realised the potential of these data, lagging behind many developed countries with respect to the availability and use of these valuable data for research for public benefit.^2^ This is due primarily to the challenges of linking data across jurisdictional boundaries and burdensome ethics and governance processes.^2^ However, recent data reforms^4–6^ have created new opportunities to build enduring linked data collections and analyse these data in more nuanced and intelligent ways to improve health outcomes in Australia and beyond.

In this context, we recently developed a novel real-world data resource, the *Medicines Intelligence (MedIntel) Data Platform*, covering over 7.4M people aged ≥18 years residing in New South Wales (NSW), Australia’s most populous state (2020 NSW adult resident population: 6.3M, approximately 32% of the Australian population^7^). The accrual period spans nearly two decades and includes data ascertained during significant events impacting the health of the Australian community, including major bushfires and the COVID-19 pandemic. This purpose-built population-based data resource has ethics and governance approval to generate high-quality, real-world, evidence across three major themes: 1) Quality use of prescribed medicines; 2) Real-world medicines safety and effectiveness; and 3) Real-world medicines costs and cost-effectiveness. Within each theme, we will focus on specific patient populations and clinical scenarios, and specific prescription medicines where contemporary evidence is lacking.

## Description of data source

### Setting

Australia has a universal healthcare system and publicly funded national health insurance scheme, known as Medicare, entitling all Australian citizens, permanent residents, and residents of countries with reciprocal health care arrangements, to a range of subsidised health services.^8^ This includes free treatment in public hospitals (funded jointly by the Federal and State/Territory governments) and subsidised treatment in private hospitals. Medicare also subsidises access to a range of outpatient services including consultations with general practitioners (GPs) and specialists. The national Pharmaceutical Benefits Scheme subsidises access to prescribed medicines dispensed in community pharmacies and private hospitals.

### Population and data sources

The *MedIntel Data Platform* is a population-based linked data resource, covering all adults (people aged ≥18 years) who were eligible for Medicare and recorded as residing in NSW at any time between 1 January 2005 and 31 December 2020. The *MedIntel Platform* comprises individual level information from four national and three NSW (State-based) data collections. For each person included in the *MedIntel* study population, records were extracted from the period 1 January 2002 (allowing a historical ascertainment period) to 31 March 2022 (most recent available at the time of data delivery). Below is a general overview of the included data collections; we provide a more detailed description in Figure 1 and Supplementary Table 1.

**Figure 1:**
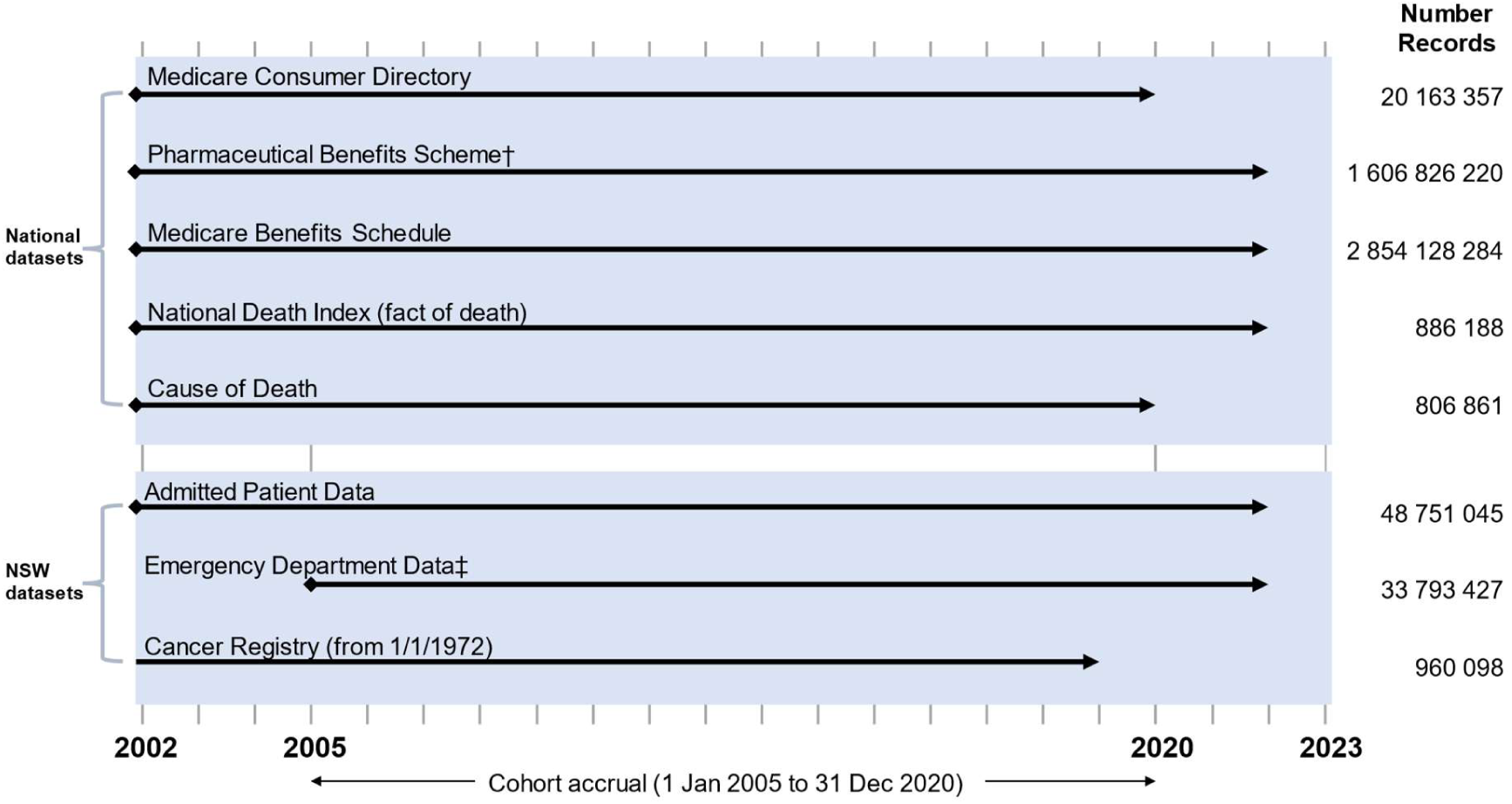
The *MedIntel Data Platform*: Data collections and time periods covered* MedIntel, Medicines Intelligence; NSW, New South Wales. The MedIntel Data Platform includes all Medicare-eligible adults (aged ≥18 years) residing in NSW at any time between 1 January 2005 and 31 December 2020. For each person included in the MedIntel study population, records were extracted from the period 1 January 2002 (allowing a historical ascertainment period) to 31 March 2022 (most recent available at the time of data delivery). * Existing approvals to update the MedIntel study population and corresponding linked datasets. † Before 1 July 2012, the Pharmaceutical Benefits Scheme data did not include records for dispensings where the price was less than the appliable government copayment. Herceptin Program data are also included from 1 Dec 2001 to 31 July 2015. ‡ EDDC records are available from 1 January 2005 onwards

#### Population: Medicare Consumer Directory (MCD)

The MCD is an administrative database of Australian citizens, permanent residents, and others eligible for Medicare,^9^ and was used to create the population spine for the *MedIntel Data Platform*.

The database contains basic demographic information for Medicare beneficiaries, including date of birth, sex, date of Medicare eligibility, and residential postcode history. We supplemented the database with an area-based index of relative socioeconomic disadvantage, derived from residential postcode and information collected in the Australian Census of Population and Housing, along with a remoteness index indicating relative road distance to services.^10,11^

#### Prescribed medicines: Pharmaceutical Benefits Scheme (PBS)

The PBS is a national program subsidising prescribed medicines listed on the basis of cost-effectiveness for specific indications in the Australian population.^12^ At June 30 2022, more than 900 unique medicines and more than 5,000 items (i.e., specific formulations, pack sizes, etc.) were PBS-listed.^13^ The PBS data collection contains dispensing records of PBS items dispensed anywhere in Australia (including details of the medicine name, strength, and Anatomical Therapeutic Chemical (ATC) five-level hierarchical classification scheme,^14^ date of supply, quantity supplied and number of repeats) that are dispensed through community pharmacies and private hospitals anywhere in Australia. For restricted medicines (i.e., those requiring additional authorisation prior to dispensing), the data also contains information on the reason for prescription (“authority code”).^15^

The Australian Federal government sets two co-payment thresholds (general and concessional), which is the maximum amount a person is required to pay for each PBS item; the government subsidises the cost of items above these thresholds. Co-payment thresholds are adjusted for inflation each calendar year. At 1 January 2024, the ‘general beneficiary’ co-payment was $AUD 31.60 for each item dispensed; concessional beneficiaries (e.g. the elderly and people on government benefits) paid $AUD 7.70.^16^ Since July 2012, PBS data have included all PBS items dispensed irrespective of patient contributions. However, before that time, only items attracting a government subsidy were captured in the data (i.e., those costing above the co-payment threshold and those dispensed to concessional beneficiaries), meaning that medicine dispensings to general beneficiaries were not fully captured in PBS data before July 2012.

The *MedIntel Data Platform* includes dispensing data from two additional publicly subsidised schemes, the Repatriation Pharmaceutical Benefits Scheme (RPBS) and the Herceptin Program. The RPBS provides subsidised access to PBS-listed medicines and additional items for Australian veterans and their dependents.^15^ The Herceptin Program was a separate publicly funded program providing subsidised access to trastuzumab for HER2+ metastatic breast cancer between December 2001 and July 2015. In July 2015, this program was closed and incorporated into the PBS.^17^ Hereafter, we refer to all three programs as the Pharmaceutical Benefits Scheme.

#### Health services: Medicare Benefits Schedule (MBS) data collection

The MBS data collection comprises records of all medical services performed anywhere in Australia that are either fully or partially subsidised by Medicare.^18^ The database includes information on consultations with physicians (i.e., general practitioners and specialists), diagnostic and therapeutic procedures (outpatient and private hospital in-patient), and pathology tests. Information on type of service provided (classified by specific MBS item codes), fees, location, registered specialty of service provider, and referring practitioner is also included.

#### Mortality: National Death Index (NDI)

The NDI contains records of all deaths occurring in Australia, supplied by the Registry of Births, Deaths and Marriages from each State or Territory.^19^ The data include date of death, residential postcode at time of death, State or Territory of death registration, and year of registration.

The NDI data also include the primary underlying cause of death and additional causes of death as recorded on each person’s death certificate. Causes of death are coded by the Australian Bureau of Statistics (ABS) using the International Classification of Diseases, Tenth Revision (ICD-10). There is generally a lag in availability of cause of death information due to the additional time required for coroner certification and information processing. Currently, cause of death data is only available to 31 December 2020.

#### Hospitalisations and diagnoses: NSW Admitted Patient Data Collection (APDC)

The APDC records information on all inpatient separations (i.e., discharges, transfers, deaths) from all public, private and repatriation hospitals, private day procedures centres, and public nursing homes in NSW.^20^ The data include information on dates of stay, diagnoses (coded according to the ICD-10 Australian Modification [ICD-10-AM]), and procedures (coded according to the Australian Classification of Health Interventions [ACHI]). Inpatient records from public and private hospitals are processed differently and there is a lag in the inclusion of private hospital records (up to one year from the date of service).

#### Emergency department visits: NSW Emergency Department Data Collection (EDDC)

The EDDC records the date, the reason for the visit (coded according to ICD Ninth Revision, Clinical Modification (ICD-9-CM); ICD-10-AM; and Systematized Medical Nomenclature for Medicine–Clinical Terminology [SNOMED CT] classifications) and the visit outcome (e.g., admission to hospital, death, or discharge) for all emergency department (ED) visits to public EDs across NSW. All larger EDs contribute data to the EDDC, covering the majority of the population.^20^ Records are available from January 2005 onwards.

#### Cancer notifications: NSW Cancer Registry (NSWCR)

The NSWCR records all invasive primary cancer cases and in-situ breast and melanoma cases diagnosed in NSW residents but excludes non-melanoma skin cancers.^20^ The registry is governed according to the International Association of Cancer Registries guidelines^21^ and records the cancer type, topography and morphology (classified according to International Classification of Diseases for Oncology - Third Edition [ICD-O-3]), date of diagnosis, degree of spread at the time of first diagnosis for solid tumours, and the date and cause of death (cancer or non-cancer) where applicable.^22^ Cancer notifications are currently available to December 2019.

### Data linkage

The Australian Institute of Health and Welfare (AIHW)^23^ and NSW Centre for Health Record Linkage (CHeReL)^24^ conducted the data linkage. The AIHW holds national-level datasets (i.e., MCD, PBS, MBS, NDI); CHeReL holds the NSW state-level collections (i.e., APDC, EDDC, NSWCR). Both the AIHW and CHeReL employ standard operating procedures in line with best practices in data linkage, including the separation principle.

Australia does not have universal person identifiers; cross-jurisdictional linkage (i.e., between national and NSW datasets) therefore requires probabilistic approaches. This process has been simplified by the establishment of jurisdiction-specific identifiers (e.g., the CHeReL Master Linkage Key [MLK], which facilitates linkage across NSW-based data collections) and an existing CHeReL-MLK-to-AIHW-identifier linkage map. These have achieved a 95% linkage rate across the jurisdictions (personal communication: *MedIntel* data linkage report).

To create the *MedIntel Data Platform*, the AIHW first identified the study population from the Medicare Consumer Directory (MCD) and assigned each person a cohort-specific Project Person Number (PPN). They then used the AIHW-to-CHeReL linkage map to identify the corresponding NSW identifiers, creating a *MedIntel*-specific CHeReL-MLK-to-PPN map. The CHeReL subsequently used the map to extract the relevant content data from the APDC, EDDC, and NSWCR, and replaced each CHeReL MLK identifier with its corresponding *MedIntel* PPN. Analogously, the AIHW extracted relevant content data from the MCD, PBS, MBS, and NDI and replaced the AIHW identifiers with the *MedIntel* PPN. The data extracts were uploaded directly by CHeReL and AIHW to a secure remote data access environment (SURE),^25^ where the linked data was made available to approved researchers for analysis (Figure 2). *No personal identifying information or jurisdiction-specific identifiers were ever made available to the researchers;* only the *MedIntel* PPNs are available, to facilitate merging across the project-specific extracts of NSW and national datasets.

**Figure 2.**
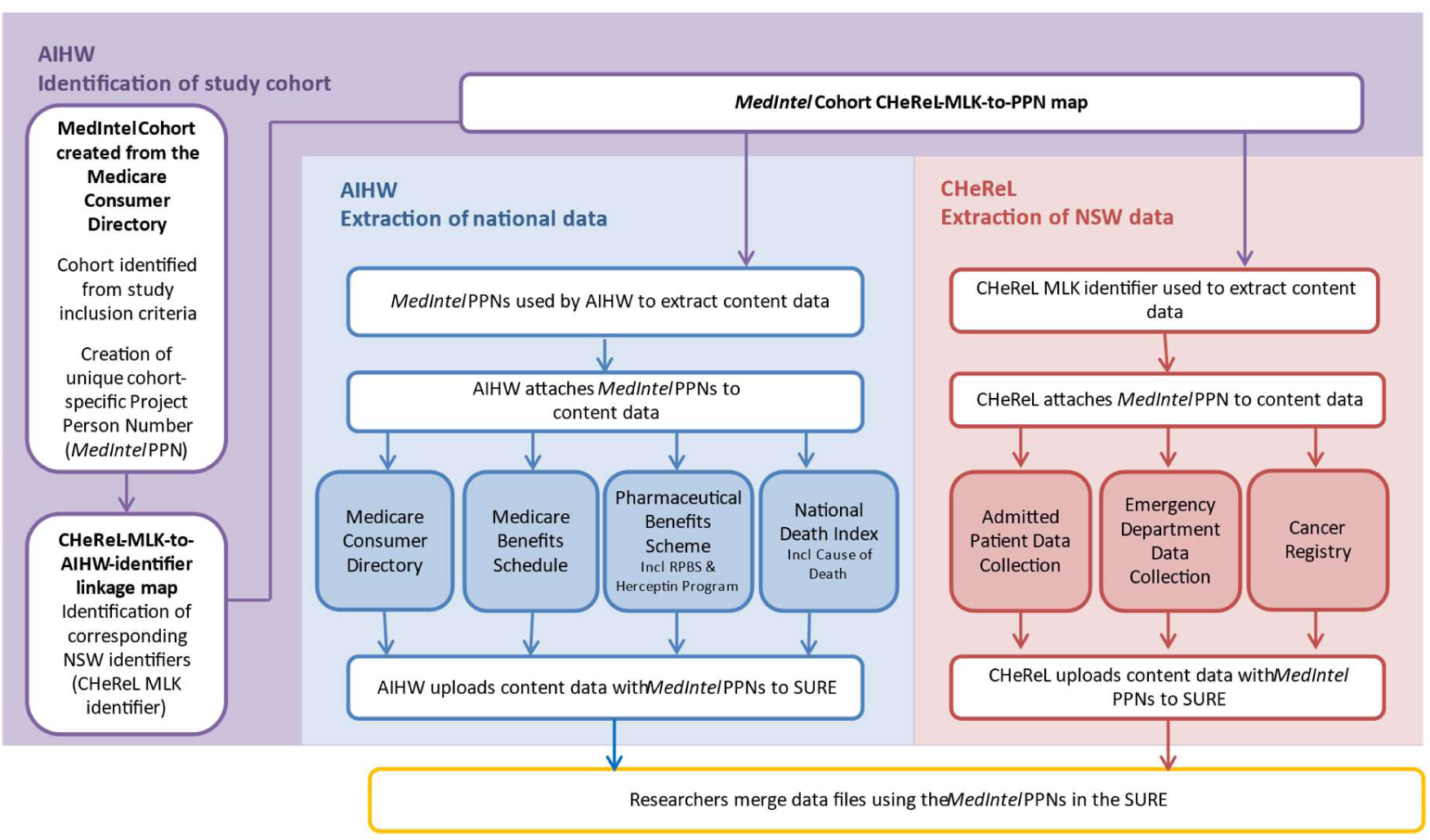
Linkage process of national and state-based data collections in *MedIntel Data Platform*, including all Medicare-eligible adult (≥18 years) residents in NSW, Australia 2005-2020 AIHW, Australian Institute of Health and Welfare; CHeReL, NSW Centre for Health Record Linkage; MedIntel, Medicines Intelligence Data Platform; MLK, Master Linkage Key; RPBS, Repatriation Pharmaceutical Benefits Scheme; SURE, Secure Unified Research Environment.

## Population characteristics and health service use

The study population includes 7.4 million unique people across all years and increased from 4.8M in 2005 to 6.0M in 2020 (Figure 3). We selected data from 2019 (the last pre-COVID-19-pandemic year) to characterise population sociodemographics (Table 1) and health service use (Table 2). Among a total of 5 980 434 persons alive on 1 January 2019, the mean age was 48.7 years and 51.1% were female. Reflecting Australia’s high degree of urbanisation, 74.7% of the population resided in a major city. The mortality rate in 2019 was 9 per 1000 or 54,003 people dying within the year. Most people had records of some health service use (Table 2); 89.4% had an MBS health service record, 73.3% a PBS medicine dispensing, and 20.5% a hospital inpatient stay during the year.

**Figure 3:**
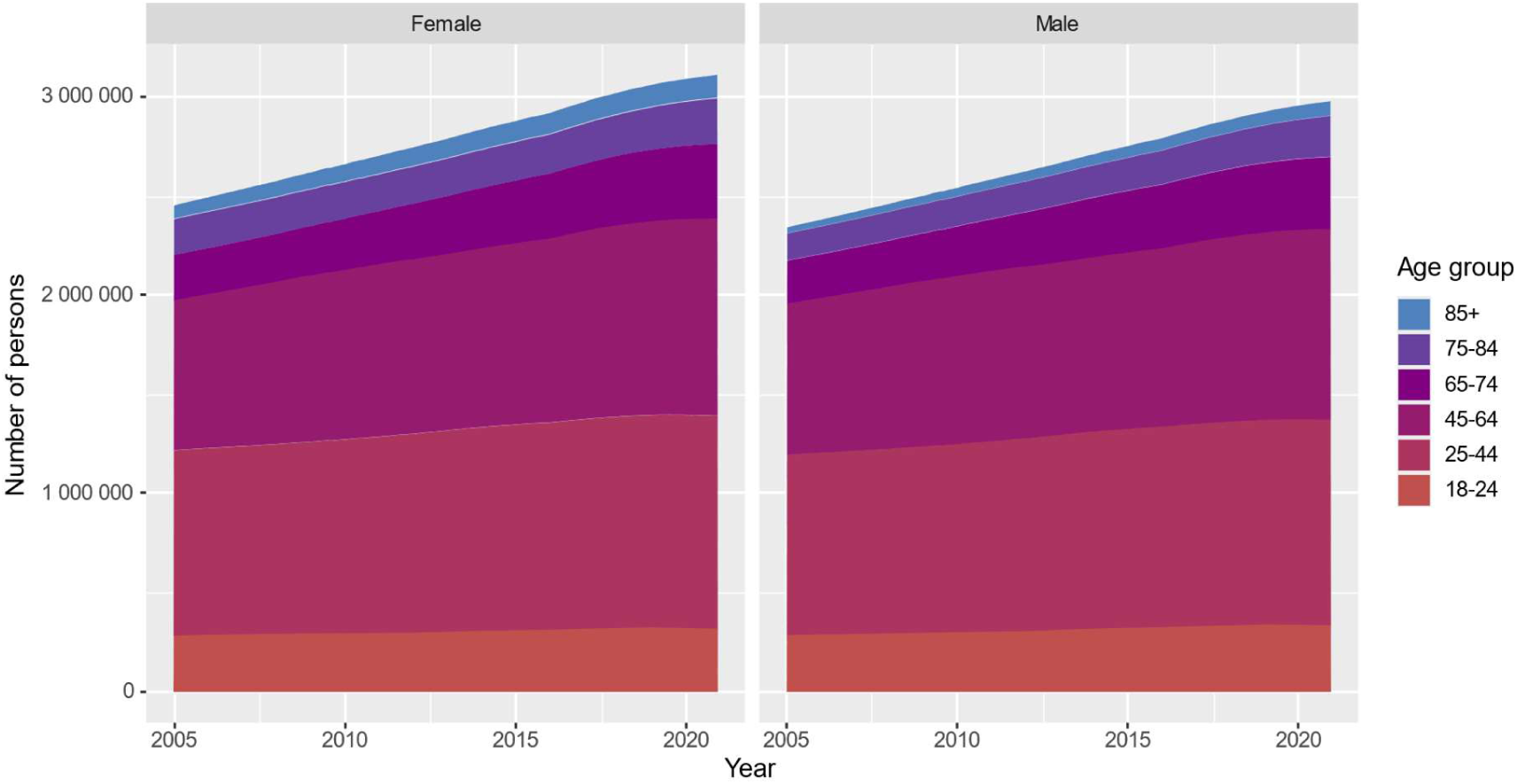
Population captured in the *MedIntel Data Platform*: Adults (≥18 years) living in NSW, Australia, 2005-2020 MedIntel, Medicines Intelligence; NSW, New South Wales.

**Table 1:** Characteristics of *MedIntel* study population as of January 1, 2019.

| Patient characteristic | Count (%) |
| --- | --- |
| **Total alive** | 5 980 434 (100%) |
| **Sex\*** |  |
| Male | 2 922 757 (48.9%) |
| Female | 3 057 677 (51.1%) |
| **Age: mean (standard deviation)** | 48.7 years (18.6) |
| **Age group** |  |
| 18–24 years | 656 292 (11.0%) |
| 25–44 years | 2 103 862 (35.2%) |
| 45–64 year | 1 921 189 (32.1%) |
| 65–74 years | 719 635 (12.0%) |
| 75–84 years | 402 884 (6.7%) |
| 85 years and older | 176 572 (3.0%) |
| **Remoteness (missing 27 588)** |  |
| Major city | 4 447 430 (74.7%) |
| Inner regional | 1 177 472 (19.8%) |
| Outer regional | 304 646 (5.1%) |
| Remote/very remote | 23 298 (0.4%) |
| **Relative socioeconomic disadvantage (missing 28 252)** |  |
| Most disadvantaged | 1 185 456 (19.9%) |
| Second quintile | 1 223 660 (20.6%) |
| Third quintile | 1 184 347 (19.9%) |
| Fourth quintile | 1 168 005 (19.6%) |
| Least disadvantaged | 1 190 714 (20.0%) |
| **Year first entered the study population** |  |
| 2005 | 4 044 470 (67.6%) |
| 2006-2010 | 725 920 (12.1%) |
| 2011-2015 | 766 696 (12.8%) |
| 2016-2019 | 443,348 (7.4%) |

MedIntel, Medicines Intelligence Data Platform
\* the only permissible values were Male and Female; other options (e.g., non-binary, other, refused to answer) were not captured in the data

**Table 2:** Prescription medicine, health service use and other health events in 2019 among people in the *MedIntel* study population as of January 1, 2019.

|  | Number of people with at least one health event (%) |  | Top 5 most common†<br>(% of total population with ≥1 record/event) |
| --- | --- | --- | --- |
| **Total population alive, NSW resident and aged ≥18 on 1 January 2019** | 5 980 434 | (100%) |  |
| **PBS medicine dispensing** | 4 382 551 | (73.3%) | Anti-infectives (43.1%), nervous system (32.2%), cardiovascular (30.2%), alimentary (26.2%), musculoskeletal (17.4%) |
| **MBS health service claims\*** | 5 349 062 | (89.4%) | GP or specialist consultation (88.8%), pathology (66.5%), diagnostic imaging (46.2%), miscellaneous (38.5%), therapeutic procedures (27.3%) |
| **Hospital in-patient stay** | 1 223 223 | (20.5%) | Primary diagnosis: Digestive system (3.6%), injury, poisoning and external causes (2.2%), neoplasms (2.2%), musculoskeletal (2.0%), circulatory (1.8%) |
| **Cancer diagnosis** | 53 862 | (0.9%) | Cancer type: Skin‡ (0.3%), urogenital (0.2%), breast (0.1%), bowel (0.1%), lymphohaematopoietic (0.1%) |
| **Emergency Department attendance** | 1 185 172 | (19.8%) | Triage category: potentially serious (9.9%), potentially life-threatening (8.9%), immediately life-threatening (4.1%), less urgent (3.0%), critically ill (0.3%) |
| **Died** | 54 003 | (0.9%) |  |

MedIntel, Medicines Intelligence Data Platform; PBS, Pharmaceutical Benefits Scheme; MBS, Medicare Benefits Schedule
† groupings were defined as: PBS, Anatomical Therapeutic Chemical (ATC) 1st level class (anatomical main group); MBS, service category; In-patient, International Statistical Classification of Diseases 10th Revision – Australian Modification (ICD-10-AM) chapter; Cancer, Australasian Association of Cancer Registries clinical cancer group; Emergency Department, NSW public hospital triage scale.
\* includes primary care, allied health, specialist, pathology and private hospital inpatient health services
‡ category includes melanoma of the skin, lip cancer, and Kaposi's sarcoma.

Figure 4 and Supplementary Tables S2-S17 demonstrate the annual prevalence of prescribed medicine use in 2019 of each ATC medicine class (first level: anatomical main group) by age group and sex. Females were more commonly dispensed medicines than males, and the prevalence of use of most medicine classes increased by age group. Overall, anti-infectives were the most prevalent medicine classes dispensed to the cohort (43.1%), followed by nervous system (32.2%) and cardiovascular system medicines (30.2%) (Table 2). Among older people, cardiovascular system medicines were most prevalent, followed by anti-infectives, nervous system medicines, and alimentary tract and metabolism medicines (Figure 4; Supplement).

**Figure 4.**
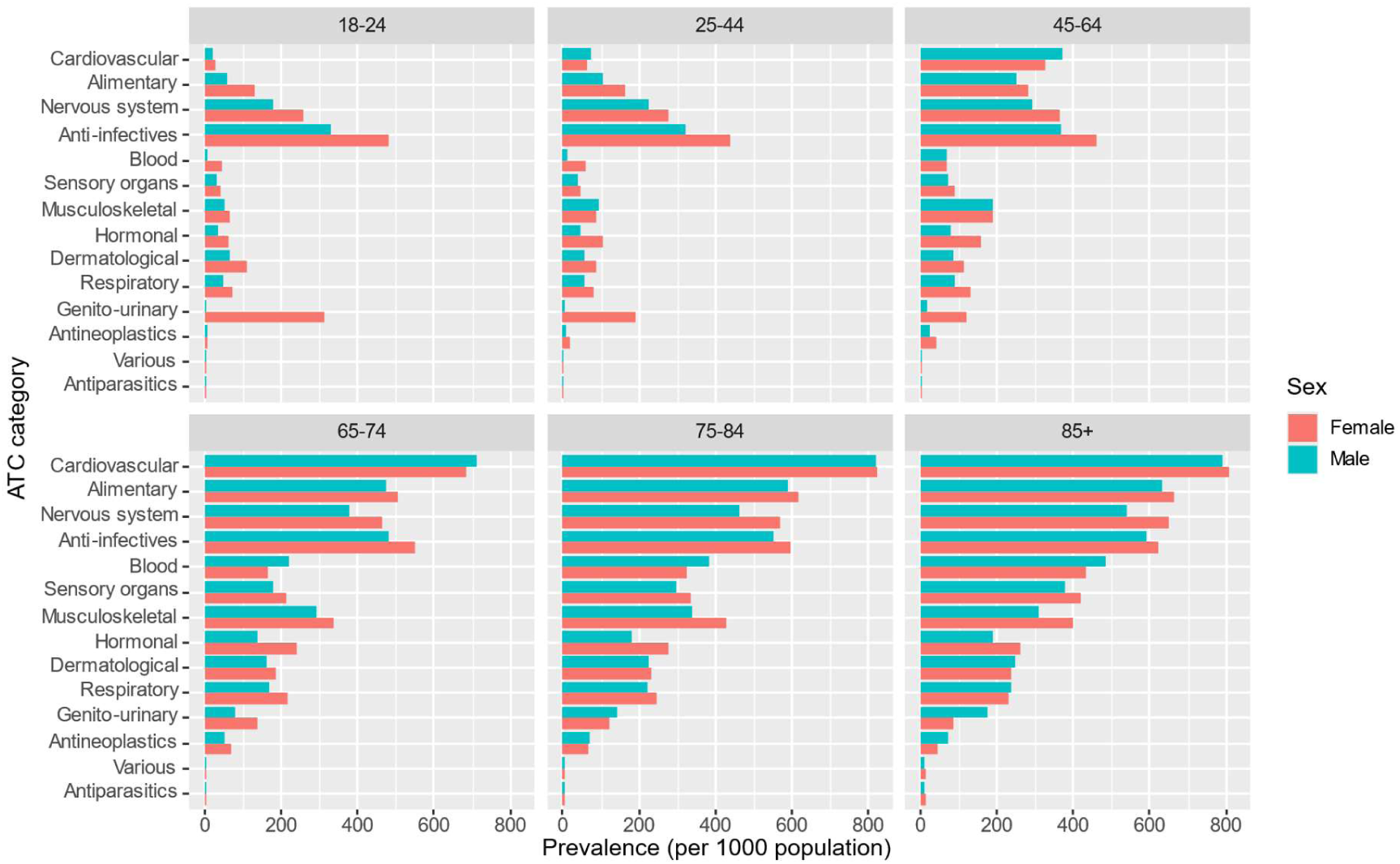
Prevalence of use (≥1 dispensing use) of each medicine class, by age and sex, among the *MedIntel* study population

## Representativeness of the *MedIntel* study population

The *MedIntel* study population represents all Medicare-eligible NSW residents aged 18 years and over, representing 36.9% of Australian adults. In 2019, the *MedIntel* cohort covered about 95% of the estimated adult resident population in NSW, with the distribution of sex, ages, remoteness of residence and socioeconomic disadvantage consistent with NSW census data.^7,10,11^ The 5% shortfall in total population numbers is due to temporary residents who are not Medicare eligible and linkage errors in the CHeReL-MLK-to-AIHW-identifier linkage map.^26^

## Strengths and limitations

The *MedIntel Data Platform* is a large, whole-of-population data collection from Australia’s most populous state that brings together comprehensive medicine exposure and outcome data. The platform contains approximately two decades of longitudinal dispensing claims, with complete capture of all PBS medicines since 2012. Outcomes are captured through data on hospitalisations, deaths, ED visits, and other health system contacts. In addition, the study period covers major health events in Australia, such as most recent bushfire season (2019 – 2020) and the COVID-19 pandemic era (2020 – 2022). The data comprising the platform are recorded for purposes of funding and reimbursement and are likely to be highly accurate descriptions of accessed health services; and international classification systems used in the datasets, such as the ATC and ICD-10 classifications, will facilitate multi-national collaborations. The *MedIntel Data Platform* captures dispensings across all major therapeutic groups, including cancer. The *MedIntel Data Platform* is a population-based resource; however, it is limited to the population of NSW, i.e. covering a whole jurisdiction but not the total nation. For residents residing near State borders, the data will lack information for hospital services accessed across State boundaries; however, all federally funded health services (medicine dispensings, Medicare services) are captured, irrespective of where in Australia they were accessed. Although the data captures biological sex at the time of Medicare registration, there is no information available on gender. Dispensing data before July 2012 solely comprise records of dispensings which attracted government subsidy, i.e., costing above the co-payment threshold. Therefore, studies examining dispensings during this time may need to restrict patient cohorts to concessional beneficiaries to ensure complete medicine capture. Similar to most other prescription databases, the PBS dispensing data do not contain information on the indication for which medicines were prescribed, but this can in some instances be deduced from the PBS item code and/or authority code associated with the dispensing.^15^ Importantly, the medicine dispensing data do not include records of medicines obtained with a private prescription (i.e., paid for out-of-pocket), dispensed to public hospital in-patients, or dispensed to patients at discharge from public hospitals. Nevertheless, it is estimated that over 90% of prescription medicine use in Australia is captured in the PBS data^27^ (including dispensing from private hospitals), and the data have been used extensively for Australian pharmacoepidemiological research.^1,2,28^ Primary care data available in the *MedIntel Data Platform* are limited to those services accessed through the MBS and only relate to the administrative record of the service, while sociodemographic data are primarily ascertained through secondary sources at area-, rather than person-level (e.g., area-level estimates of social advantage/disadvantage ascertained via Census). Finally, the *MedIntel Data Platform* does not contain complete immigration/emigration data, but movement between NSW and other States and Territories can be detected based on patient residence postcode data.

## Plans and perspectives

The first supply of data for the *MedIntel Platform* was delivered in November 2022. The next data update is planned for late 2024, at which time we will seek approval to link additional data collections to further advance the capability of the collection to explore medicine use and outcomes. We are discussing potential linkage with custodians of longitudinal cohort data, including self-reported medical and behavioural data, as well as with the Australian Bureau of Statistics who hold health, social, and Census data. These data will allow us to control for confounders more robustly (e.g., individual-level socioeconomic and health status, medical history, ethnicity, etc.) and to monitor medicine use in specific sub-populations.

We currently have project-specific approvals for studies across therapeutic areas such as cancer, mental health, pain, and cardiometabolic conditions, funded by Australian and international research grants.

## Data access

The *MedIntel Data Platform* was developed using best-practice governance and access criteria and we welcome national and international research collaborations. Our website^29^ details the specific requirements for researchers to undertake projects using the platform.

## Ethics and data storage

The MedIntel Data Platform and associated research program on medicine use, safety, effectiveness, costs and cost-effectiveness has ethical approval from: AIHW Human Research Ethics Committee (AIHW HREC; approval number EO2021/1/1233) and NSW Population and Health Services Research Ethics Committee (PHSREC; approval number 2020/ETH02273). Individual research projects conducted under this ethical approval require that a study-specific protocol be approved by PHSREC and additional personnel be approved by AIHW. Our HREC approvals permit annual updates of the study population and additional years of data for each person already included in the study population.

The data platform is housed in the Secure Unified Research Environment (SURE), managed by the Sax Institute, a trusted, independent third party.^25^ SURE is a safe setting, offering data controls meeting the highest data governance and security requirements. Housing the data in SURE adheres to the Five Safes framework^30–32^—safe projects, people, settings, data and outputs. All analyses must be conducted within SURE.

## Data Availability

The *MedIntel Data Platform* was developed using best-practice governance and access criteria and we welcome national and international research collaborations. Specific requirements for researchers wishing to undertake projects using the platform are available on our website (https://unsw.to/medinteldp)

## Acknowledgments

The authors would like to thank the Australian Government Department of Health and Aged Care and the New South Wales (NSW) Ministry of Health for data provision and the Australian Institute of Health and Welfare (AIHW) and NSW Centre for Health Record Linkage (CHeReL) for undertaking the data linkage process. Secure data access was provided through the Sax Institute’s Secure Unified Research Environment (SURE).

## Author contributions

SAP, HZ, ML acquired the data. ML, MBG, XC prepared the data. SAP, HZ, MOF, MBG, ML, XC interpreted the data and drafted the manuscript. All authors critically reviewed and approved the final manuscript.

