## Supplementary Materials for "The Medicines Intelligence Data Platform: A population-based data resource from New South Wales, Australia"

‡ joint first authors

**Supplementary Table S1.** Data collections and key measures captured in the *MedIntel Data Platform*

**Supplementary Table S2.** Prevalence of use (≥1 dispensing) of each medicine class in 2019, by sex, among the *MedIntel* study population aged 18-24 years

**Supplementary Table S3.** Prevalence of use (≥1 dispensing) of each medicine class in 2019, by sex, among the *MedIntel* study population aged 25-29 years

**Supplementary Table S4.** Prevalence of use (≥1 dispensing) of each medicine class in 2019, by sex, among the *MedIntel* study population aged 30-34 years

**Supplementary Table S5.** Prevalence of use (≥1 dispensing) of each medicine class in 2019, by sex, among the *MedIntel* study population aged 35-39 years

**Supplementary Table S6.** Prevalence of use (≥1 dispensing) of each medicine class in 2019, by sex, among the *MedIntel* study population aged 40-44 years

**Supplementary Table S7.** Prevalence of use (≥1 dispensing) of each medicine class in 2019, by sex, among the *MedIntel* study population aged 45-49 years

**Supplementary Table S8.** Prevalence of use (≥1 dispensing) of each medicine class in 2019, by sex, among the *MedIntel* study population aged 50-54 years

**Supplementary Table S9.** Prevalence of use (≥1 dispensing) of each medicine class in 2019, by sex, among the *MedIntel* study population aged 55-59 years

**Supplementary Table S10.** Prevalence of use (≥1 dispensing) of each medicine class in 2019, by sex, among the *MedIntel* study population aged 60-64 years

**Supplementary Table S11.** Prevalence of use (≥1 dispensing) of each medicine class in 2019, by sex, among the *MedIntel* study population aged 65-69 years

**Supplementary Table S12.** Prevalence of use (≥1 dispensing) of each medicine class in 2019, by sex, among the *MedIntel* study population aged 70-74 years

**Supplementary Table S13.** Prevalence of use (≥1 dispensing) of each medicine class in 2019, by sex, among the *MedIntel* study population aged 75-79 years

**Supplementary Table S14.** Prevalence of use (≥1 dispensing) of each medicine class in 2019, by sex, among the *MedIntel* study population aged 80-84 years

**Supplementary Table S15.** Prevalence of use (≥1 dispensing) of each medicine class in 2019, by sex, among the *MedIntel* study population aged 85-89 years

**Supplementary Table S16.** Prevalence of use (≥1 dispensing) of each medicine class in 2019, by sex, among the *MedIntel* study population aged 90-94 years

**Supplementary Table S17.** Prevalence of use (≥1 dispensing) of each medicine class in 2019, by sex, among the *MedIntel* study population aged 95 years and older

**Supplementary Table S1.** Data collections and key measures captured in the *MedIntel Data Platform*

| | Data collection | Key variables |
|---|---|---|
| National data collections | **Medicare Consumer Directory (MCD):** includes individual-level records for all Australian citizens, permanent residents and others eligible for Australia's universal healthcare system, known as Medicare. | • Date of birth (month and year)<br>• Sex<br>• Start and end date of postcode of residence<br>• Residential postcode history<br>• Residential remoteness (based on postcode and Census)<br>• Index of relative socioeconomic disadvantage (based on postcode and Census) |
| | **Pharmaceutical Benefits Scheme (PBS)[1]:** includes individual-level claim records of all[a] PBS items[b] dispensed at community pharmacies and private hospitals across Australia. | • Date of medicine dispensing and prescribing<br>• PBS item code indicating medicine and strength (mapped to ATC class)<br>• Dispensed quantity and number of scripts filled<br>• Amount subsidised and amount paid by patient<br>• Concessional (social security beneficiary) status<br>• Authority code<br>• Prescriber specialty and postcode<br>• Pharmacy type and location |
| | **Medicare Benefits Schedule (MBS)[2]:** includes individual-level claim records for medical services covered by government subsidy. | • Date of service and referral for service<br>• Item code indicating service provided<br>• Type of service provided (e.g., professional service, diagnostic procedures, imaging)<br>• Amount paid by patient, amount reimbursed by Medicare<br>• Hospital flag (service delivered in hospital or not)<br>• Provider and referral practitioner location and specialty |
| | **National Death Index (NDI)[3]:** includes individual-level records of all deaths occurring and registered in Australia. | • Date of death<br>• Underlying and other causes of death (ICD-10 classification) – only available until 31 Dec 2020<br>• Postcode of residence at death<br>• State and year of death registration |
| State-based data collections (NSW)[4] | **NSW Admitted Patient Data Collection (APDC):** includes individual-level claim records for all inpatient separations (i.e., discharges, transfers, deaths) from all public, private and repatriation hospitals, private day procedures centres and public nursing homes in NSW. | • Date of admission, date and type of separation<br>• Diagnoses and causes of injury (ICD-10-AM classification)<br>• Procedures performed (ACHI classification)<br>• Length of stay<br>• Referral and discharge status<br>• Other demographics[c]: country of birth and residence, marital status<br>• Public / private hospital flag<br>• Facility type and location (local health district) |
| | **NSW Emergency Department Data Collection (EDDC)**: includes individual-level claim records for all emergency department visits to approximately 90 public emergency departments across NSW. | • Date of presentation and departure<br>• Diagnoses (ICD-9-CM, ICD-10-AM, or SNOMED-CT classifications)<br>• Triage categories and visit type<br>• Status (mode of separation) and referral on discharge<br>• Other demographics: country of birth and marital status<br>• Facility type and location (local health district) |
| | **NSW Cancer Registry (NSWCR):** includes individual level case records for all invasive cancer cases (excluding non-melanoma skin cancers) and in-situ melanoma and breast cancer cases diagnosed in NSW residents. | • Date of cancer diagnosis (month and year)<br>• Cancer type, site, topography and morphology at diagnosis (ICD9, ICD10 and WHO ICD-O-3 classification)<br>• Degree of spread at diagnosis and basis (method) for diagnosis |

| | | |
|---|---|---|
| | | • Cancer death details<br>• Other demographics: country of birth, remoteness and socioeconomic position (based on residence at diagnosis) |

ACHI, Australian Classification of Health Interventions; ATC, Anatomical Therapeutic Chemical classification; ICD-9-CM, International Classification of Diseases 9th Revision, Clinical Modification; ICD-10, International Classification of Diseases 10th Revision; ICD-10-AM, International Classification of Diseases 10th Revision, Australian Modification; ICD-O-3, International Classification of Diseases for Oncology, 3rd Edition; MedIntel, Medicines Intelligence; NSW, New South Wales; SNOMED-CT, Systematized Nomenclature of Medicine – Clinical Terms

[a] Before July 2012, medicines costing below the patient co-payment amount are not captured in the data

[b] Also includes dispensing data from two additional publicly subsidised schemes, the Repatriation Pharmaceutical Benefits Scheme (RPBS) and The Herceptin Program

[c] Refers to demographic data other than date of birth, sex and location of residence.

**Supplementary Table S2.** Prevalence of use (≥1 dispensing) of each medicine class in 2019, by sex, among the *MedIntel* study population aged 18-24 years

| Sex | 1st level ATC class | Rate per 1000 population |
|---|---|---|
| Females | Alimentary | 129.0 |
| | Blood | 45.0 |
| | Cardiovascular | 27.9 |
| | Dermatological | 109.4 |
| | Genito-urinary | 314.3 |
| | Hormonal | 63.6 |
| | Anti-infectives | 482.6 |
| | Antineoplastics | 7.1 |
| | Musculoskeletal | 63.8 |
| | Nervous system | 259.0 |
| | Antiparasitics | 2.3 |
| | Respiratory | 73.4 |
| | Sensory organs | 42.0 |
| | Various | 0.4 |
| Males | Alimentary | 59.6 |
| | Blood | 6.0 |
| | Cardiovascular | 19.8 |
| | Dermatological | 66.0 |
| | Genito-urinary | 3.8 |
| | Hormonal | 34.6 |
| | Anti-infectives | 329.7 |
| | Antineoplastics | 5.9 |
| | Musculoskeletal | 50.5 |
| | Nervous system | 180.2 |
| | Antiparasitics | 1.6 |
| | Respiratory | 48.7 |
| | Sensory organs | 31.4 |
| | Various | 0.4 |

MedIntel, Medicines Intelligence data platform

**Supplementary Table S3.** Prevalence of use (≥1 dispensing) of each medicine class in 2019, by sex, among the *MedIntel* study population aged 25-29 years

| Sex | 1st level ATC class | Rate per 1000 population |
|---|---|---|
| Females | Alimentary | 158.7 |
| | Blood | 55.4 |
| | Cardiovascular | 35.1 |
| | Dermatological | 91.4 |
| | Genito-urinary | 244.2 |
| | Hormonal | 81.4 |
| | Anti-infectives | 458.4 |
| | Antineoplastics | 13.3 |
| | Musculoskeletal | 69.4 |
| | Nervous system | 272.7 |
| | Antiparasitics | 2.0 |
| | Respiratory | 73.0 |
| | Sensory organs | 44.0 |
| | Various | 0.4 |
| Males | Alimentary | 76.0 |
| | Blood | 7.5 |
| | Cardiovascular | 28.3 |
| | Dermatological | 54.2 |
| | Genito-urinary | 3.7 |
| | Hormonal | 38.3 |
| | Anti-infectives | 313.3 |
| | Antineoplastics | 7.5 |
| | Musculoskeletal | 69.7 |
| | Nervous system | 210.5 |
| | Antiparasitics | 1.3 |
| | Respiratory | 49.6 |
| | Sensory organs | 32.6 |
| | Various | 0.5 |

MedIntel, Medicines Intelligence data platform

**Supplementary Table S4.** Prevalence of use (≥1 dispensing) of each medicine class in 2019, by sex, among the *MedIntel* study population aged 30-34 years

| Sex | 1st level ATC class | Rate per 1000 population |
|---|---|---|
| Females | Alimentary | 163.1 |
| | Blood | 62.5 |
| | Cardiovascular | 43.3 |
| | Dermatological | 85.0 |
| | Genito-urinary | 196.3 |
| | Hormonal | 102.2 |
| | Anti-infectives | 441.4 |
| | Antineoplastics | 17.2 |
| | Musculoskeletal | 74.2 |
| | Nervous system | 257.7 |
| | Antiparasitics | 2.0 |
| | Respiratory | 73.9 |
| | Sensory organs | 45.3 |
| | Various | 0.5 |
| Males | Alimentary | 91.4 |
| | Blood | 10.1 |
| | Cardiovascular | 47.2 |
| | Dermatological | 53.8 |
| | Genito-urinary | 3.7 |
| | Hormonal | 45.5 |
| | Anti-infectives | 317.6 |
| | Antineoplastics | 8.8 |
| | Musculoskeletal | 84.4 |
| | Nervous system | 211.9 |
| | Antiparasitics | 1.3 |
| | Respiratory | 52.8 |
| | Sensory organs | 37.4 |
| | Various | 0.5 |

MedIntel, Medicines Intelligence data platform

**Supplementary Table S5.** Prevalence of use (≥1 dispensing) of each medicine class in 2019, by sex, among the *MedIntel* study population aged 35-39 years

| Sex | 1st level ATC class | Rate per 1000 population |
|---|---|---|
| Females | Alimentary | 160.4 |
| | Blood | 62.3 |
| | Cardiovascular | 63.7 |
| | Dermatological | 84.7 |
| | Genito-urinary | 176.6 |
| | Hormonal | 115.4 |
| | Anti-infectives | 431.0 |
| | Antineoplastics | 19.7 |
| | Musculoskeletal | 89.4 |
| | Nervous system | 272.8 |
| | Antiparasitics | 2.1 |
| | Respiratory | 82.3 |
| | Sensory organs | 48.0 |
| | Various | 0.6 |
| Males | Alimentary | 111.4 |
| | Blood | 14.1 |
| | Cardiovascular | 81.5 |
| | Dermatological | 56.2 |
| | Genito-urinary | 4.2 |
| | Hormonal | 51.4 |
| | Anti-infectives | 328.8 |
| | Antineoplastics | 9.8 |
| | Musculoskeletal | 101.9 |
| | Nervous system | 225.9 |
| | Antiparasitics | 1.4 |
| | Respiratory | 58.5 |
| | Sensory organs | 42.1 |
| | Various | 0.6 |

MedIntel, Medicines Intelligence data platform

**Supplementary Table S6.** Prevalence of use (≥1 dispensing) of each medicine class in 2019, by sex, among the *MedIntel* study population aged 40-44 years

| Sex | 1st level ATC class | Rate per 1000 population |
|---|---|---|
| Females | Alimentary | 170.9 |
| | Blood | 64.4 |
| | Cardiovascular | 106.3 |
| | Dermatological | 87.6 |
| | Genito-urinary | 154.6 |
| | Hormonal | 117.4 |
| | Anti-infectives | 418.6 |
| | Antineoplastics | 23.4 |
| | Musculoskeletal | 111.4 |
| | Nervous system | 303.9 |
| | Antiparasitics | 2.1 |
| | Respiratory | 95.1 |
| | Sensory organs | 51.5 |
| | Various | 0.9 |
| Males | Alimentary | 134.7 |
| | Blood | 20.2 |
| | Cardiovascular | 136.9 |
| | Dermatological | 59.8 |
| | Genito-urinary | 5.3 |
| | Hormonal | 56.2 |
| | Anti-infectives | 326.7 |
| | Antineoplastics | 11.9 |
| | Musculoskeletal | 123.1 |
| | Nervous system | 248.3 |
| | Antiparasitics | 1.4 |
| | Respiratory | 65.0 |
| | Sensory organs | 44.6 |
| | Various | 0.8 |

MedIntel, Medicines Intelligence data platform

**Supplementary Table S7.** Prevalence of use (≥1 dispensing) of each medicine class in 2019, by sex, among the *MedIntel* study population aged 45-49 years

| Sex | 1st level ATC class | Rate per 1000 population |
|---|---|---|
| Females | Alimentary | 204.9 |
| | Blood | 65.4 |
| | Cardiovascular | 175.1 |
| | Dermatological | 94.4 |
| | Genito-urinary | 134.9 |
| | Hormonal | 123.9 |
| | Anti-infectives | 422.2 |
| | Antineoplastics | 30.4 |
| | Musculoskeletal | 139.8 |
| | Nervous system | 338.4 |
| | Antiparasitics | 2.3 |
| | Respiratory | 105.6 |
| | Sensory organs | 60.8 |
| | Various | 1.1 |
| Males | Alimentary | 170.7 |
| | Blood | 31.5 |
| | Cardiovascular | 215.7 |
| | Dermatological | 64.1 |
| | Genito-urinary | 7.2 |
| | Hormonal | 61.3 |
| | Anti-infectives | 329.9 |
| | Antineoplastics | 14.9 |
| | Musculoskeletal | 145.4 |
| | Nervous system | 266.8 |
| | Antiparasitics | 1.7 |
| | Respiratory | 69.2 |
| | Sensory organs | 50.8 |
| | Various | 1.2 |

MedIntel, Medicines Intelligence data platform

**Supplementary Table S8.** Prevalence of use (≥1 dispensing) of each medicine class in 2019, by sex, among the *MedIntel* study population aged 50-54 years

| Sex | 1st level ATC class | Rate per 1000 population |
|---|---|---|
| Females | Alimentary | 249.6 |
| | Blood | 57.8 |
| | Cardiovascular | 274.0 |
| | Dermatological | 105.3 |
| | Genito-urinary | 114.8 |
| | Hormonal | 145.3 |
| | Anti-infectives | 442.8 |
| | Antineoplastics | 40.0 |
| | Musculoskeletal | 170.0 |
| | Nervous system | 356.0 |
| | Antiparasitics | 2.3 |
| | Respiratory | 118.5 |
| | Sensory organs | 76.3 |
| | Various | 1.4 |
| Males | Alimentary | 222.4 |
| | Blood | 51.0 |
| | Cardiovascular | 321.0 |
| | Dermatological | 77.2 |
| | Genito-urinary | 11.7 |
| | Hormonal | 71.7 |
| | Anti-infectives | 354.6 |
| | Antineoplastics | 19.4 |
| | Musculoskeletal | 176.1 |
| | Nervous system | 282.5 |
| | Antiparasitics | 1.9 |
| | Respiratory | 81.5 |
| | Sensory organs | 61.7 |
| | Various | 1.8 |

MedIntel, Medicines Intelligence data platform

**Supplementary Table S9.** Prevalence of use (≥1 dispensing) of each medicine class in 2019, by sex, among the *MedIntel* study population aged 55-59 years

| Sex | 1st level ATC class | Rate per 1000 population |
|---|---|---|
| Females | Alimentary | 310.6 |
| | Blood | 61.5 |
| | Cardiovascular | 388.2 |
| | Dermatological | 122.1 |
| | Genito-urinary | 112.9 |
| | Hormonal | 172.5 |
| | Anti-infectives | 474.3 |
| | Antineoplastics | 47.2 |
| | Musculoskeletal | 203.4 |
| | Nervous system | 375.7 |
| | Antiparasitics | 2.7 |
| | Respiratory | 141.4 |
| | Sensory organs | 97.7 |
| | Various | 1.9 |
| Males | Alimentary | 282.9 |
| | Blood | 78.8 |
| | Cardiovascular | 437.1 |
| | Dermatological | 92.7 |
| | Genito-urinary | 20.3 |
| | Hormonal | 86.2 |
| | Anti-infectives | 383.9 |
| | Antineoplastics | 25.9 |
| | Musculoskeletal | 207.2 |
| | Nervous system | 304.2 |
| | Antiparasitics | 2.2 |
| | Respiratory | 99.7 |
| | Sensory organs | 79.5 |
| | Various | 2.5 |

MedIntel, Medicines Intelligence data platform

**Supplementary Table S10.** Prevalence of use (≥1 dispensing) of each medicine class in 2019, by sex, among the *MedIntel* study population aged 60-64 years

| Sex | 1st level ATC class | Rate per 1000 population |
|---|---|---|
| Females | Alimentary | 386.8 |
| | Blood | 87.9 |
| | Cardiovascular | 512.4 |
| | Dermatological | 142.9 |
| | Genito-urinary | 121.1 |
| | Hormonal | 199.0 |
| | Anti-infectives | 508.7 |
| | Antineoplastics | 55.4 |
| | Musculoskeletal | 250.2 |
| | Nervous system | 400.6 |
| | Antiparasitics | 2.8 |
| | Respiratory | 171.5 |
| | Sensory organs | 131.9 |
| | Various | 2.5 |
| Males | Alimentary | 354.9 |
| | Blood | 119.2 |
| | Cardiovascular | 557.9 |
| | Dermatological | 113.6 |
| | Genito-urinary | 35.3 |
| | Hormonal | 103.6 |
| | Anti-infectives | 418.6 |
| | Antineoplastics | 35.0 |
| | Musculoskeletal | 241.4 |
| | Nervous system | 325.3 |
| | Antiparasitics | 2.7 |
| | Respiratory | 120.7 |
| | Sensory organs | 105.2 |
| | Various | 3.0 |

MedIntel, Medicines Intelligence data platform

**Supplementary Table S11.** Prevalence of use (≥1 dispensing) of each medicine class in 2019, by sex, among the *MedIntel* study population aged 65-69 years

| Sex | 1st level ATC class | Rate per 1000 population |
|---|---|---|
| Females | Alimentary | 469.5 |
| | Blood | 136.3 |
| | Cardiovascular | 637.0 |
| | Dermatological | 173.4 |
| | Genito-urinary | 135.6 |
| | Hormonal | 230.1 |
| | Anti-infectives | 538.6 |
| | Antineoplastics | 63.8 |
| | Musculoskeletal | 303.9 |
| | Nervous system | 439.9 |
| | Antiparasitics | 3.3 |
| | Respiratory | 206.0 |
| | Sensory organs | 184.0 |
| | Various | 2.9 |
| Males | Alimentary | 439.4 |
| | Blood | 185.0 |
| | Cardiovascular | 673.8 |
| | Dermatological | 146.6 |
| | Genito-urinary | 63.3 |
| | Hormonal | 125.7 |
| | Anti-infectives | 465.1 |
| | Antineoplastics | 46.0 |
| | Musculoskeletal | 275.4 |
| | Nervous system | 356.0 |
| | Antiparasitics | 3.5 |
| | Respiratory | 153.8 |
| | Sensory organs | 152.5 |
| | Various | 4.1 |

MedIntel, Medicines Intelligence data platform

**Supplementary Table S12.** Prevalence of use (≥1 dispensing) of each medicine class in 2019, by sex, among the *MedIntel* study population aged 70-74 years

| Sex | 1st level ATC class | Rate per 1000 population |
|---|---|---|
| Females | Alimentary | 544.4 |
| | Blood | 201.6 |
| | Cardiovascular | 737.7 |
| | Dermatological | 201.5 |
| | Genito-urinary | 136.6 |
| | Hormonal | 256.2 |
| | Anti-infectives | 563.9 |
| | Antineoplastics | 71.2 |
| | Musculoskeletal | 372.5 |
| | Nervous system | 490.7 |
| | Antiparasitics | 3.9 |
| | Respiratory | 231.0 |
| | Sensory organs | 246.0 |
| | Various | 3.6 |
| Males | Alimentary | 515.7 |
| | Blood | 263.0 |
| | Cardiovascular | 756.3 |
| | Dermatological | 181.6 |
| | Genito-urinary | 99.7 |
| | Hormonal | 149.3 |
| | Anti-infectives | 503.0 |
| | Antineoplastics | 59.1 |
| | Musculoskeletal | 314.2 |
| | Nervous system | 400.3 |
| | Antiparasitics | 4.1 |
| | Respiratory | 185.5 |
| | Sensory organs | 209.8 |
| | Various | 5.4 |

MedIntel, Medicines Intelligence data platform

**Supplementary Table S13.** Prevalence of use (≥1 dispensing) of each medicine class in 2019, by sex, among the *MedIntel* study population aged 75-79 years

| Sex | 1st level ATC class | Rate per 1000 population |
|---|---|---|
| Females | Alimentary | 602.6 |
| | Blood | 286.2 |
| | Cardiovascular | 808.7 |
| | Dermatological | 227.5 |
| | Genito-urinary | 129.5 |
| | Hormonal | 273.5 |
| | Anti-infectives | 587.4 |
| | Antineoplastics | 69.8 |
| | Musculoskeletal | 417.2 |
| | Nervous system | 546.2 |
| | Antiparasitics | 5.7 |
| | Respiratory | 248.9 |
| | Sensory organs | 312.1 |
| | Various | 4.5 |
| Males | Alimentary | 573.4 |
| | Blood | 352.8 |
| | Cardiovascular | 811.0 |
| | Dermatological | 216.1 |
| | Genito-urinary | 132.1 |
| | Hormonal | 173.5 |
| | Anti-infectives | 539.5 |
| | Antineoplastics | 68.4 |
| | Musculoskeletal | 336.8 |
| | Nervous system | 442.3 |
| | Antiparasitics | 5.4 |
| | Respiratory | 215.8 |
| | Sensory organs | 275.5 |
| | Various | 6.0 |

MedIntel, Medicines Intelligence data platform

**Supplementary Table S14.** Prevalence of use (≥1 dispensing) of each medicine class in 2019, by sex, among the *MedIntel* study population aged 80-84 years

| Sex | 1st level ATC class | Rate per 1000 population |
|---|---|---|
| Females | Alimentary | 638.2 |
| | Blood | 373.4 |
| | Cardiovascular | 842.4 |
| | Dermatological | 241.4 |
| | Genito-urinary | 116.3 |
| | Hormonal | 282.9 |
| | Anti-infectives | 606.5 |
| | Antineoplastics | 60.2 |
| | Musculoskeletal | 441.7 |
| | Nervous system | 596.1 |
| | Antiparasitics | 6.7 |
| | Respiratory | 244.2 |
| | Sensory organs | 362.7 |
| | Various | 5.1 |
| Males | Alimentary | 614.0 |
| | Blood | 430.1 |
| | Cardiovascular | 830.9 |
| | Dermatological | 235.4 |
| | Genito-urinary | 161.2 |
| | Hormonal | 190.8 |
| | Anti-infectives | 569.4 |
| | Antineoplastics | 74.1 |
| | Musculoskeletal | 344.3 |
| | Nervous system | 493.5 |
| | Antiparasitics | 7.1 |
| | Respiratory | 230.1 |
| | Sensory organs | 330.3 |
| | Various | 6.8 |

MedIntel, Medicines Intelligence data platform

**Supplementary Table S15.** Prevalence of use (≥1 dispensing) of each medicine class in 2019, by sex, among the *MedIntel* study population aged 85-89 years

| Sex | 1st level ATC class | Rate per 1000 population |
|---|---|---|
| Females | Alimentary | 663.5 |
| | Blood | 437.5 |
| | Cardiovascular | 839.5 |
| | Dermatological | 241.6 |
| | Genito-urinary | 98.0 |
| | Hormonal | 276.8 |
| | Anti-infectives | 623.0 |
| | Antineoplastics | 49.1 |
| | Musculoskeletal | 433.5 |
| | Nervous system | 641.3 |
| | Antiparasitics | 10.8 |
| | Respiratory | 239.2 |
| | Sensory organs | 404.6 |
| | Various | 8.9 |
| Males | Alimentary | 635.4 |
| | Blood | 487.7 |
| | Cardiovascular | 818.6 |
| | Dermatological | 249.5 |
| | Genito-urinary | 175.2 |
| | Hormonal | 196.9 |
| | Anti-infectives | 593.3 |
| | Antineoplastics | 73.5 |
| | Musculoskeletal | 329.3 |
| | Nervous system | 531.0 |
| | Antiparasitics | 10.0 |
| | Respiratory | 238.3 |
| | Sensory organs | 368.1 |
| | Various | 7.5 |

MedIntel, Medicines Intelligence data platform

**Supplementary Table S16.** Prevalence of use (≥1 dispensing) of each medicine class in 2019, by sex, among the *MedIntel* study population aged 90-94 years

| Sex | 1st level ATC class | Rate per 1000 population |
|---|---|---|
| Females | Alimentary | 678.1 |
| | Blood | 454.6 |
| | Cardiovascular | 798.1 |
| | Dermatological | 236.0 |
| | Genito-urinary | 78.7 |
| | Hormonal | 258.7 |
| | Anti-infectives | 638.7 |
| | Antineoplastics | 39.5 |
| | Musculoskeletal | 378.5 |
| | Nervous system | 678.0 |
| | Antiparasitics | 13.8 |
| | Respiratory | 234.2 |
| | Sensory organs | 442.1 |
| | Various | 18.7 |
| Males | Alimentary | 645.5 |
| | Blood | 503.0 |
| | Cardiovascular | 776.4 |
| | Dermatological | 251.9 |
| | Genito-urinary | 180.6 |
| | Hormonal | 186.2 |
| | Anti-infectives | 606.5 |
| | Antineoplastics | 72.5 |
| | Musculoskeletal | 292.6 |
| | Nervous system | 569.5 |
| | Antiparasitics | 12.5 |
| | Respiratory | 242.8 |
| | Sensory organs | 400.7 |
| | Various | 15.6 |

MedIntel, Medicines Intelligence data platform

**Supplementary Table S17.** Prevalence of use (≥1 dispensing) of each medicine class in 2019, by sex, among the *MedIntel* study population aged 95 years and older

| Sex | 1st level ATC class | Rate per 1000 population |
|---|---|---|
| Females | Alimentary | 614.9 |
| | Blood | 369.3 |
| | Cardiovascular | 674.6 |
| | Dermatological | 205.6 |
| | Genito-urinary | 53.4 |
| | Hormonal | 204.5 |
| | Anti-infectives | 580.4 |
| | Antineoplastics | 26.1 |
| | Musculoskeletal | 267.1 |
| | Nervous system | 627.8 |
| | Antiparasitics | 15.5 |
| | Respiratory | 190.3 |
| | Sensory organs | 436.0 |
| | Various | 20.8 |
| Males | Alimentary | 543.0 |
| | Blood | 389.7 |
| | Cardiovascular | 575.8 |
| | Dermatological | 222.0 |
| | Genito-urinary | 146.5 |
| | Hormonal | 142.8 |
| | Anti-infectives | 501.9 |
| | Antineoplastics | 54.7 |
| | Musculoskeletal | 203.2 |
| | Nervous system | 496.0 |
| | Antiparasitics | 11.0 |
| | Respiratory | 207.5 |
| | Sensory organs | 371.4 |
| | Various | 33.7 |

MedIntel, Medicines Intelligence data platform
